# Mathematical Modelling of Oxygenation Dynamics using High-Resolution Perfusion Data – Part 2: Physiological Insights

**DOI:** 10.1101/2024.11.12.24317213

**Authors:** Mansour T. A. Sharabiani, Alireza S. Mahani, Richard W. Issitt, Yadav Srinivasan, Alex Bottle, Serban Stoica

## Abstract

**Background:** Accurate prediction of oxygen demand is essential for optimizing oxygen delivery during paediatric cardiopulmonary bypass (CPB), but traditional models may not fully account for the influences of temperature, age, and weight. We aimed to quantify these relationships and assess age-related variability in the oxygen extraction ratio (OER) response.

**Methods:** We analysed data from 334 paediatric patients undergoing CPB, developing an extended GARIX model (eGARIX) that incorporates age, weight, and nonparametric temperature modelling via splines. Subgroup analyses compared OER responses to changes in haemoglobin and arterial oxygen saturation across different age groups.

**Results:** eGARIX significantly improved model fit over GARIX (p < 0.001). Oxygen demand per body surface area exhibited a nonlinear relationship with age and weight, peaking around 3 years. In neonates and infants, oxygen demand positively correlated with weight; in adolescents, the correlation was negative. The temperature dependence was more complex than constant-Q_10_ models predict, showing reduced sensitivity of oxygen demand to mild hypothermia and increased sensitivity at deep hypothermia. Younger patients showed a diminished OER response to haemoglobin changes.

**Conclusions:** Age and weight significantly affect oxygen demand during paediatric CPB, highlighting the need for individualized models. Our findings challenge constant-Q_10_ assumptions, indicating that nonparametric models better capture temperature effects. Personalized oxygen delivery strategies are essential; further validation is recommended.

## Introduction

Accurately predicting oxygen demand is critical for determining optimal oxygen delivery levels in paediatric patients during cardiopulmonary bypass (CPB). However, real-time prediction remains challenging due to its complex dependence on multiple physiological factors, including body temperature and patient characteristics.

In Part 1 of this research series [1], we developed the GARIX model to predict minute-by-minute changes in the oxygen extraction ratio (OER) during paediatric CPB surgeries, capturing the adaptive OER response to fluctuations in oxygen demand driven by intraoperative temperature changes.

In this paper, we explore the physiological implications of GARIX, addressing two limitations of the baseline model. First, although our data cover paediatric patients up to 18 years old, they span a wide age range with potentially significant physiological differences. To account for this heterogeneity, we include patient age and weight in the steady-state component of the model, allowing oxygen demand parameters to vary with these attributes. We also divide the data into four age groups to compare the dynamic coefficients of eGARIX models trained within each age group.

Second, the baseline model assumes that oxygen demand depends on temperature according to the van’t Hoff equation, implying a constant Q_10_ (the multiplicative increase in oxygen demand per 10°C increase). However, evidence for this assumption is limited, based on only two temperature data points from previous studies [2–6]. We relax this assumption and empirically model the temperature dependence using a nonparametric spline fit.

Additionally, we demonstrate how the fitted eGARIX model can simulate the Fox82 experiments, showing that the hyperbolic dependence of VO₂i on DO_2_i observed by Fox82 can emerge from a piecewise-linear model combined with insufficient wait times between state transitions. We also illustrate how over-oxygenation can occur due to the slow adaptation of OER to changes in variables such as cardiac flow.

Our aim is to deepen the understanding of the physiological determinants of oxygen demand in paediatric patients by constructing a more nuanced model that accounts for key variables like age, weight, and temperature, thereby contributing to the development of personalized oxygen delivery strategies during paediatric CPB.

For a list of nonstandard abbreviations and acronyms used in this manuscript, see Supplementary Material A.

## Patients and Methods

### Study Design and Participants

Cohort is a retrospective analysis of consecutive patients aged younger than 18 years (maximum age treated at the institution) undergoing 963 CPB surgeries at Great Ormond Street Hospital between 2019 and 2021. Clinical data were collated in a research platform within the hospital’s governance structure and de-identified before analysis. Institutional approval was given for the undertaking of this project (audit number 3045). Individual consent was not required because only routinely collected de-identified hospital data were evaluated within a secure digital research environment as part of an existing research database approval (17/LO/0008).

Details on Data Collection, Anaesthesia, and CPB are provided in Supplementary Material B.

### Data Preparation

Data preparation and analysis were conducted using R 4.4.0 and Python 3.11.7. For baseline data preparation, refer to Part 1. We analysed age both as a continuous variable and categorically, defining age groups as: neonates (<1 month), infants (1–6 months), toddlers (6 months–3 years), and children (>3 years), with a maximum age of 18.23 years.

Consistent with Part 1, we excluded patients who developed postoperative acute kidney injury (AKI) from training GARIX models to focus on physiological responses, resulting in a dataset of 334 patients undergoing 343 CPBs, totalling 19,687 minutes of operation time. AKI was defined according to KDIGO criteria [9]. There are two exceptions to this rule. First is when comparing our model to Fox82. Since the experiments conducted by Fox82 did not explicitly screen for ‘no-AKI’, we included all our patients, regardless of AKI status, in this comparison to maximise comparability. Second is in creating mean response surface and conditional/marginal means for oxygen demand vs. age/weight, where we used the entire patient population to enhance the statistical power of the empirical representation of the joint density of age and weight in children. Our total patient population is 963, of which 926 have valid age and weight values.

### GARIX (Baseline Model)

The baseline GARIX model, introduced in Part 1, predicts minute-by-minute changes in OER during paediatric CPB, assuming changes in logit-transformed OER follow i.i.d. normal distributions.

The expected change in logit(OER) at each time point comprises three components:

1. **Equilibrium Term Group (ETG)**: Corrective force to balance oxygen consumption with temperature-dependent oxygen demand.
2. **Autoregressive Term Group (ATG)**: Captures influence of past OER changes.
3. **Exogenous Term Group (XTG)**: Accounts for effects of current and past changes in exogenous variables, including cardiac index (CI), haemoglobin concentration (Hb), arterial oxygen saturation (SaO_2_), and temperature (Temp).

We use a lag term of n=7 minutes, as determined optimal in Part 1.

For detailed mathematical formulation and model diagnostics, see Part 1.

### eGARIX (Extended GARIX)

We extend the baseline model by incorporating patient age, weight, their interaction, and modelling temperature dependence using a spline:

Baseline ETG (Part 1):

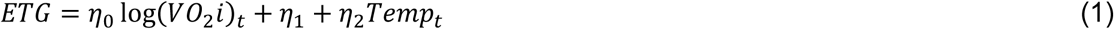

where

- *η*_0_, *η*_1_, *η*_2_: coefficients,
- *VO*_2_*i*_*t*_: Indexed oxygen consumption at time t,
- *Temp*_*t*_: Temperature at time t.

Extended ETG:

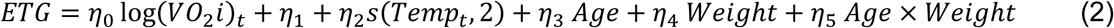

Here, *s*(*Temp*, 2) is a spline function of *Temp* with two degrees of freedom, capturing nonlinear temperature dependence. *η*_3_, *η*_4_, *η*_4_ are coefficients of age, weight and their interaction, respectively.

At steady state (ETG = 0), oxygen consumption equals oxygen demand, yielding:

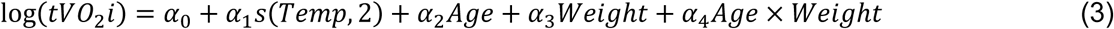

Where 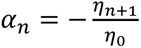, *n* = 0,1,2,3,4. *tVO*_2_*i* is indexed target oxygen consumption (or oxygen demand).

*Q*_10_ is calculated as:

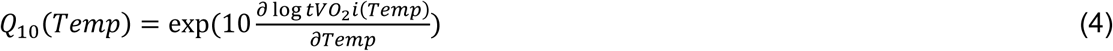

Under van’t Hoff specification, 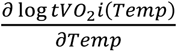 (partial derivative of oxygen demand with respect to temperature) is constant, making *Q*_10_ independent of *Temp*. We used finite differences to approximate Equation 4 for plotting *Q*_10_ versus *Temp*.

Note that, in producing Figures 5 (comparison with Fox82) and 6 (oxygenation gap), we used GARIX rather than eGARIX, since the focus there is not on studying the effects of age, weight or nonlinear dependence of oxygen demand on temperature.

### Schofield Equations

The Schofield equations [8] estimate basal metabolic rate (BMR) in kcal/day as a linear function of weight (kg), with coefficients varying by age and gender, based on a large dataset of indirect calorimetry measurements. They are commonly used to predict energy expenditure across diverse populations.

#### For males

- Age 0–3 years: BMR = 59.512 Weight – 30.4
- Age 3–10 years: BMR = 22.706 Weight + 504.3
- Age 10–18 years: BMR = 17.686 Weight + 658.2

#### For females

- Age 0–3 years: BMR = 58.317 Weight − 31.1
- Age 3–10 years: BMR = 20.315 Weight + 485.9
- Age 10–18 years: BMR = 13.384 Weight + 692.6

To compare eGARIX predictions of oxygen demand, in mL/min, to Schofield equations for metabolic rate, in kcal/day, we apply a conversion factor of 7.2 to the former, based on the established assumption that 5 kcal of energy is expended per litre of oxygen consumed [15].

### Mean Response Surface and Conditional/Marginal Means

The mean response surface for oxygen demand as a function of age and weight is formed empirically by 1) holding the temperature at a pre-specified value, and 2) evaluating the fitted eGARIX (or Schofield) equations for the entire data (including patients with AKI). (Schofield equations are valid for normal body temperature of 37°C.) Conditional means are formed by holding one of the two variables (age or weight) fixed and varying the other. For marginal means, we fit a LOESS (locally estimated scatterplot smoothing) curve to the pairs of (oxygen demand, age/weight) that form the mean response surface.

### Consistency Assessment

<describe how consistency of eGARIX vs. Schofield curves was calculated>

To assess consistency of metabolic rate predictions by eGARIX and Schofield, we calculated the variance of the difference between the logarithms of the two numbers evaluated at each prediction point. F test was used to assess the statistical significance of the difference in variance between models.

## Results

### Patients and Outcomes

Detailed patient characteristics and outcomes are identical to those in Part 1 [1].

### Model Performance

Adding the interaction term between age and weight significantly improved upon GARIX (coefficient −5.5 × 10^-2^ for predicting log *VO*_2_*i*, p = 0.008). The spline term for temperature was also significant (ANOVA p < 0.001). Likelihood-ratio tests confirmed that eGARIX significantly improved over GARIX (p < 0.001).

### Comparison to Schofield

Figure 1 compares the predicted metabolic rate vs. age for eGARIX and Schofield models. Note that the two curves are nearly parallel. Considering that the y axis is in logarithmic scale, this implies a near-fixed ratio between metabolic rate predictions from the two models (average ratio: 0.54). We also note that eGARIX produced the most consistent curve with Schofield, compared to GARIX as well as the static model (with or without age and weight terms). For instance, the variance of differences between log predictions from (baseline) GARIX and Schofield is 3.8 times the same figure for eGARIX vs. Schofield (p-value < 0.001). Note that this is independent of the conversion factor used to map oxygen demand to metabolic rate (see Methods).

**Figure 1.**
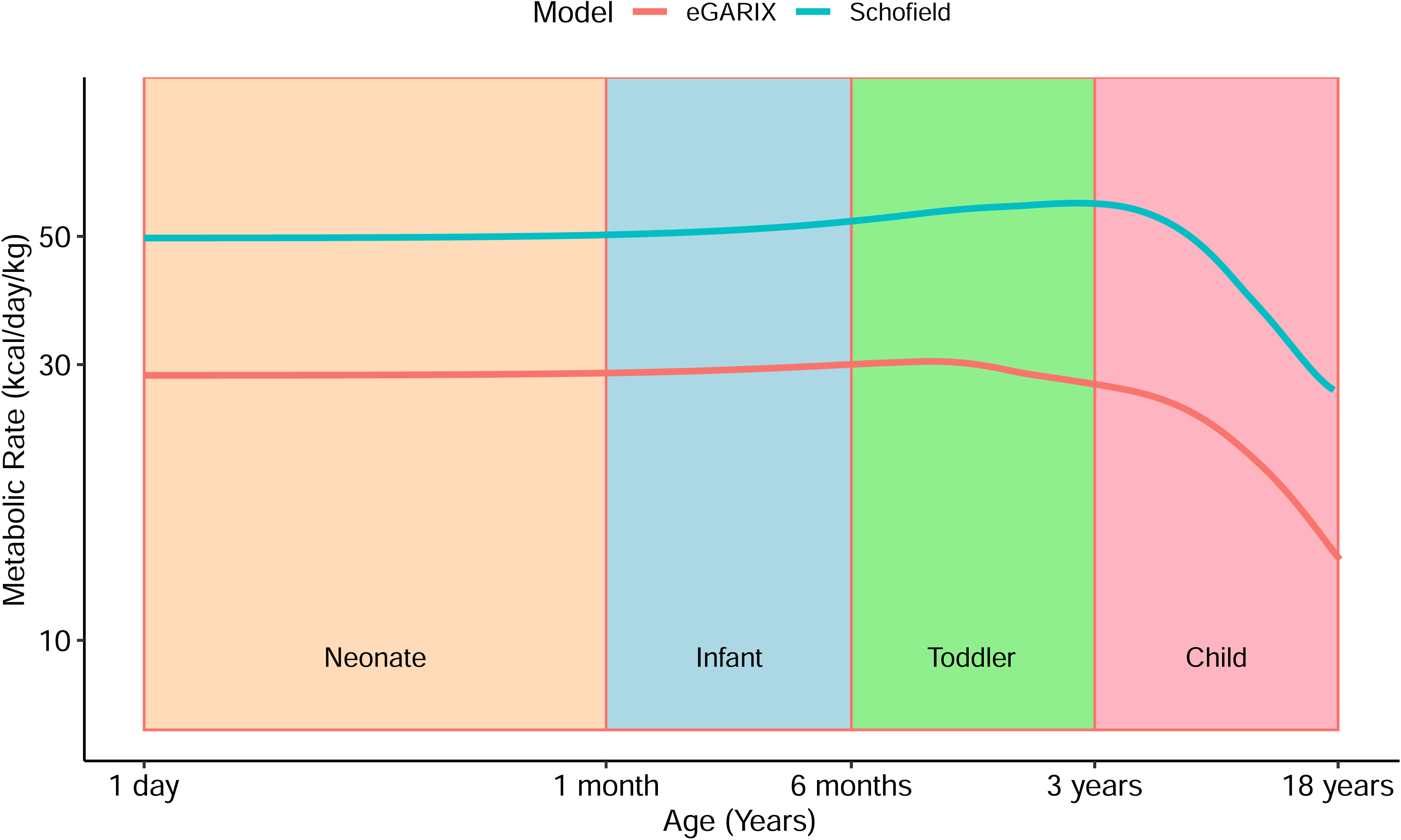
Metabolic Rate vs. Age. Comparison of predicted metabolic rate – per kilogram of body weight – vs. age between Schofield and eGARIX. Each data point corresponds to a patient in our dataset (including AKI). LOESS fits to each dataset are overlaid. Note that x and y axes are displayed in log scale.

### Age and Weight Effects on Oxygen Demand and OER Dynamics

Figure 2A illustrates the predicted oxygen demand (tVO_2_i) as a function of age and weight. In younger patients, oxygen demand positively correlates with weight, but this relationship weakens and reverses in older children due to the negative interaction term between age and weight. Panel B shows that a 10% increase in weight leads to a 2% increase in oxygen demand in neonates but a 1% decrease in 8-year-olds. Projections onto the age–tVO_2_i and weight–tVO_2_i planes (Figures 2C and 2D) reveal that oxygen demand increases with age, peaking at approximately 85 mL/min/m² at 3 years, then declining to around 70 mL/min/m² in teenagers and as low as 42 mL/min/m² in neonates.

**Figure 2.**
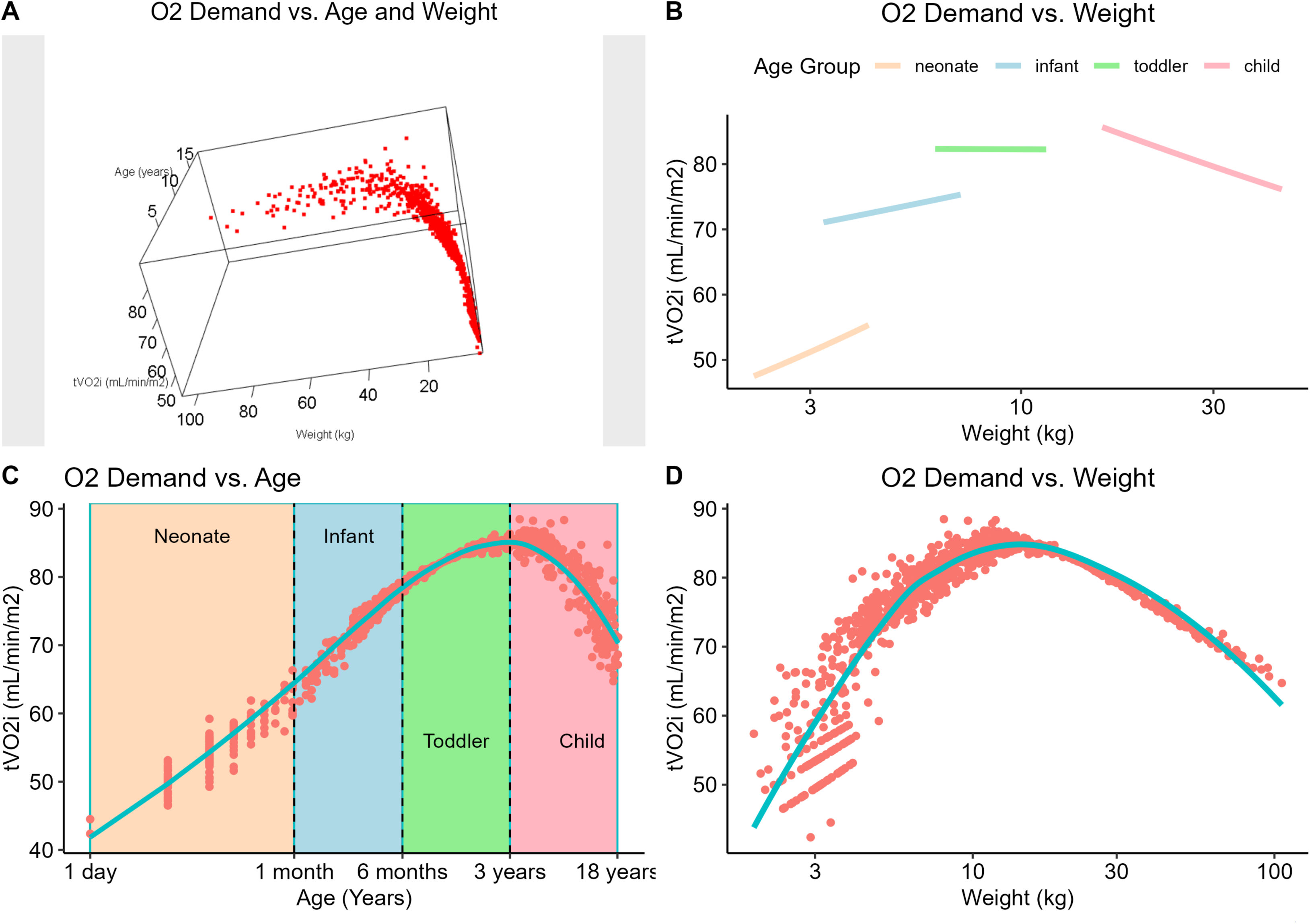
Oxygen Demand vs. Patient Age and Weight. A) Oxygen demand (*tVO*_2_*i*) vs patient age and weight, evaluated at (age, weight) pairs for patients in our data (including AKI). B) Oxygen demand vs. weight at four different ages each representing an age group (neonate: 5 days, infant: 3 months, toddler: 1 year, child: 8 years). All oxygen demand numbers are calculated at a temperature of 35C using the eGARIX (7) model. C) Projection of points in A onto the age-*tVO*_2_*i* plane. Green line is a nonparametric (loess) regression of *tVO*_2_*i* on age. Colour-shared areas represent neonate (orange), infant (blue), toddler (green) and child (pink) age groups. D) Projection of points in A onto the weight-*tVO*_2_*i* plane.

Figure 3 displays the coefficients for the exogenous terms log(Hb) and log(SaO_2_) across different lags and age groups. Younger patients show a weaker OER response to changes in haemoglobin but a more pronounced immediate response to changes in arterial oxygen saturation compared to older children. Supplementary Material C shows that other state variables do not exhibit clear age-related trends.

**Figure 3.**
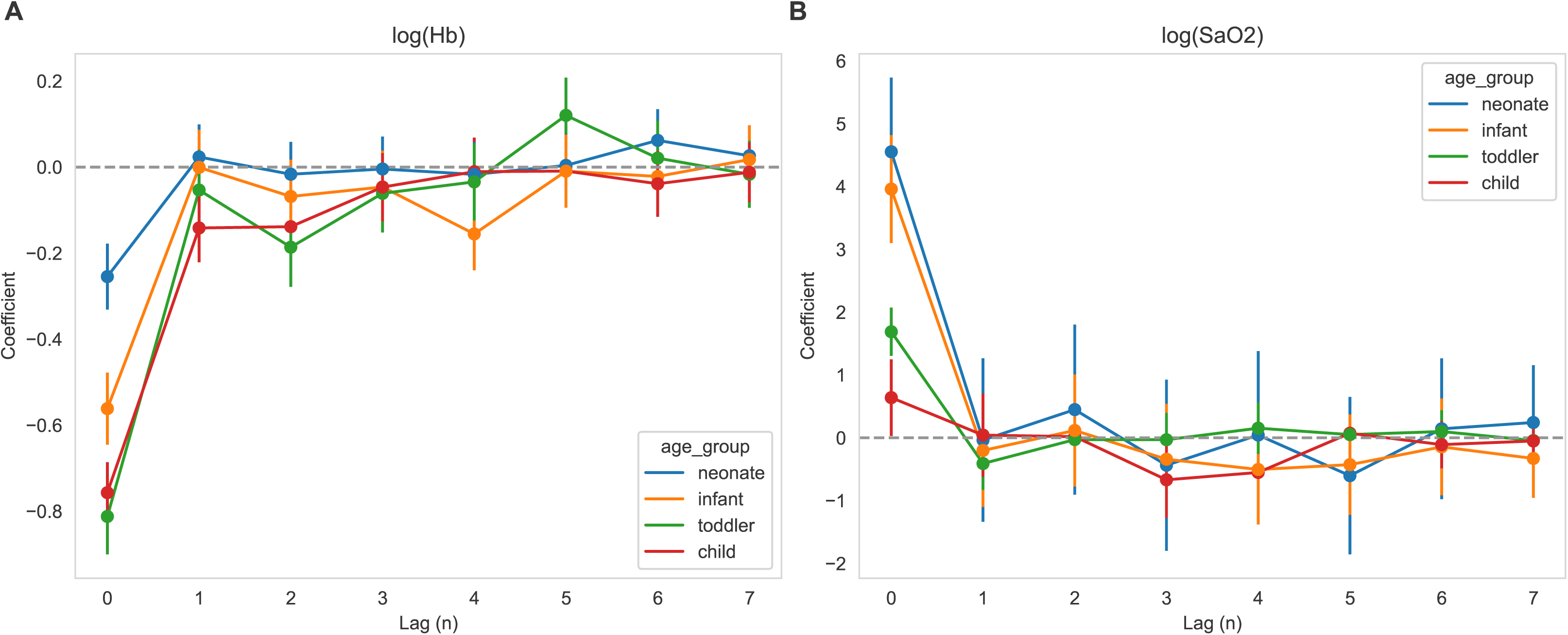
OER Dynamics by Age group. Coefficients of change terms in eGARIX involving haemoglobin (log *Hb*, left) and arterial oxygen saturation (log *SaO*_2_, right), as a function of lag (n). Each colour corresponds to an eGARIX(7) model that is trained on a subset of patients in a single age group. See Methods for exact definitions of change terms, lag and age groups.

### Oxygen Demand vs. Temperature

Figure 4A shows the geometrically averaged oxygen demand – as predicted by eGARIX – versus temperature, while Figure 4B depicts Q_10_ as a function of temperature. Q_10_ decreases from 3.8 at 16°C to 1.1 at 35°C, indicating a nonlinear relationship. Specifically, reducing temperature from 36.8°C to 31.8°C leads to only an 11% reduction in oxygen demand, whereas the next 5°C drop results in a 37% reduction. In contrast, GARIX predicts a constant Q_10_ of 2.2, implying a consistent 33% reduction per 5°C decrease. The wider confidence intervals at low temperatures reflect the sparsity of data below 25°C (only 4% of data points).

**Figure 4.**
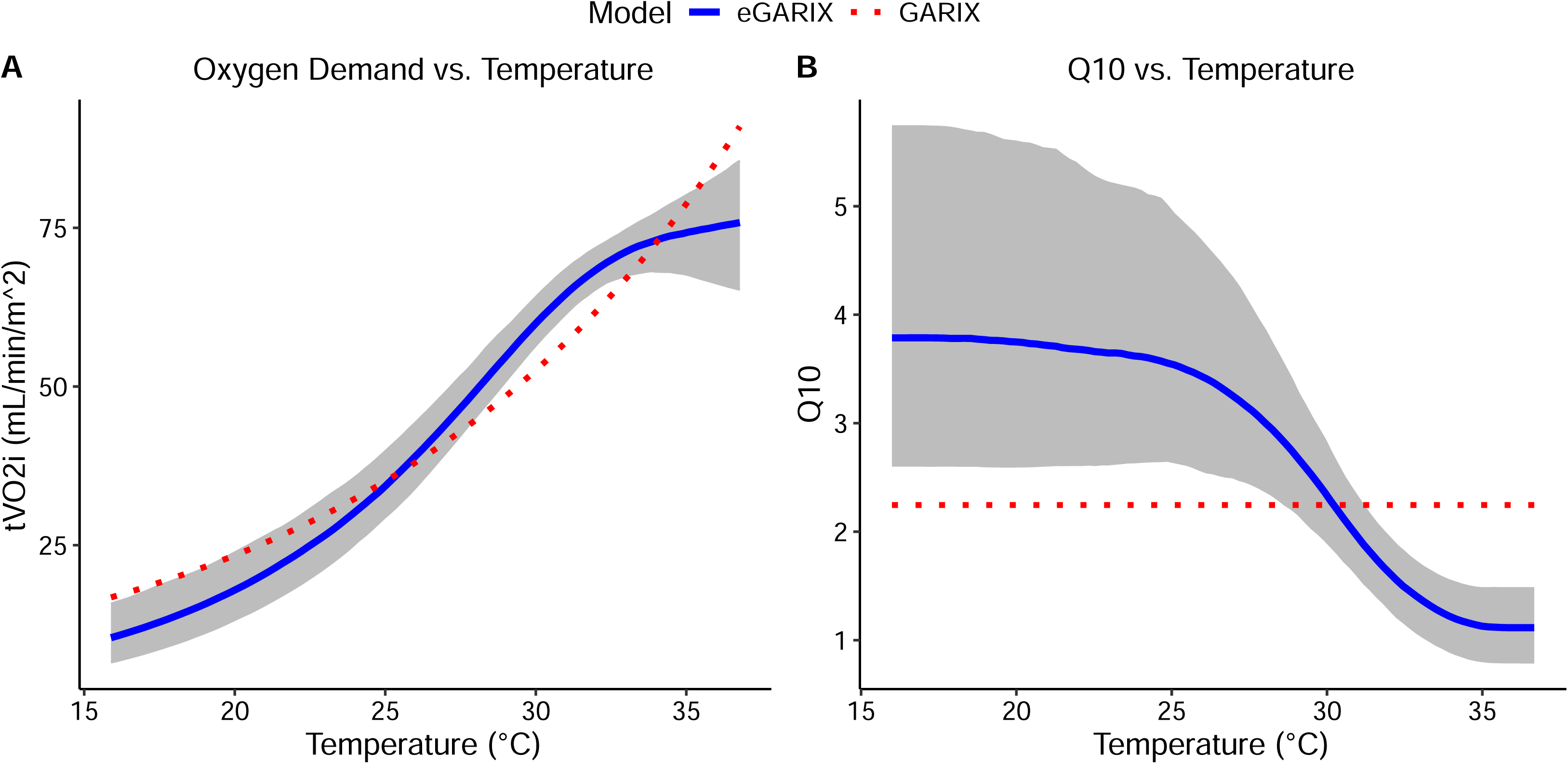
Oxygen Demand vs. Temperature. Oxygen Demand vs. Temperature. A: Estimated oxygen demand vs. temperature, with each point being the geometric average of the predicted values (by eGARIX(7)) for the unique (age, weight) combinations in training data. B: Estimated Q10 vs. temperature, with each point being the arithmetic average of Q10 values for each (age, weight) pair in the data. Grey regions represent 95% confidence bands, using 1000 bootstrapped samples. Dashed red lines represent predictions from the GARIX(7) (baseline) model.

### GARIX vs. Fox82

We simulated the Fox82 human experiments using GARIX (Supplementary Material D). With a 10-minute wait time between cardiac index (CI) changes, the GARIX+ results closely resemble Fox82’s VO_2_i versus CI scatterplot (Figure 5), and our hyperbolic fit parameters align within the 95% confidence intervals of Fox82’s estimates (Supplementary Material D, Table D.1). However, since GARIX+ predicts adaptation times longer than 10 minutes, increasing the wait time to 100 minutes yields a fit closer to the piecewise linear assumption, approaching it asymptotically with longer waits.

**Figure 5.**
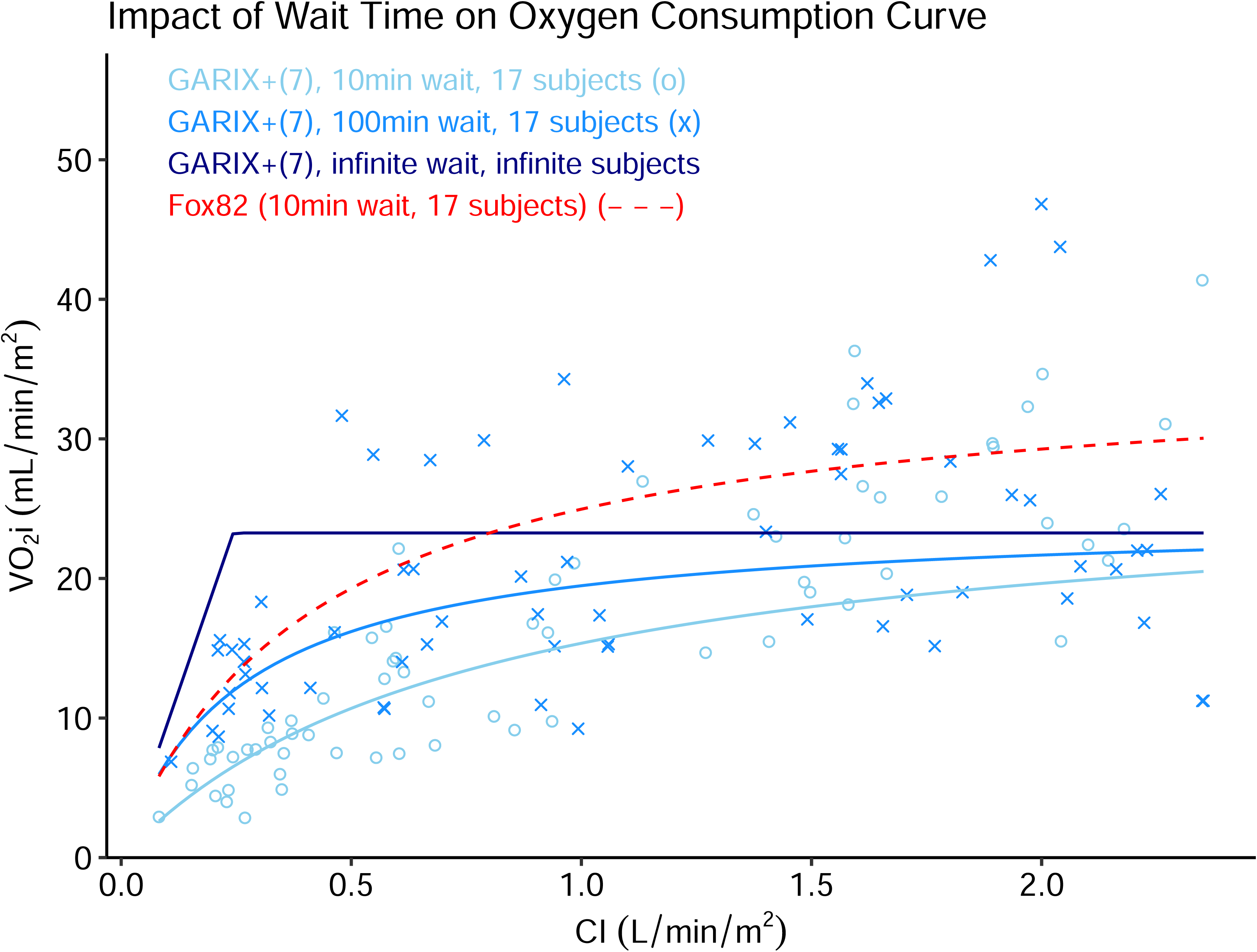
GARIX vs. Fox82. Comparison of VO2i vs. CI curves, produced by GARIX (simulation-based) and the hyperbolic model of Fox82 (human experiments). Light blue ‘o’ marks show results of a sample stochastic simulation using GARIX(7), with settings that closely match Fox82 (see Supplementary Material D), including 17 subjects and a 10-minute wait after each step change to the CI. Light blue curve is the hyperbolic fit to simulated data. Blue ‘x’ marks are GARIX+(7) stochastic simulations using wait times of 100 minutes. The blue curve shows the hyperbolic fit to that data. The dark blue, piecewise linear line shows the results for infinite wait time and averaging over an infinite pool of (simulated) subjects. Dotted red curve is the hyperbolic fit of Fox82 to their human experiments.

Kirklin and Blackstone [2] combined Fox82’s human data at 20°C with 1950s animal data at 37°C [4–6], using the van’t Hoff model to interpolate oxygen consumption across temperatures (Table 1, "Kirklin – Asymptotic"). We compared these with eGARIX predictions at various ages and temperatures. Although not directly comparable due to differences in subject age and species, we observe that eGARIX predictions (for children) and "Kirklin – Clinical" values are relatively close in the 20-30°C but diverge at normal temperatures, with Kirklin implying a significantly higher oxygen demand.

**Table 1:** eGARIX vs. Kirklin – Oxygen Demand and Temperature. Comparison of Kirklin and GARIX/eGARIX predictions for oxygen demand at different temperatures. ‘Kirklin – Asymptotic’ corresponds to oxygen consumption at infinite cardiac index using the hyperbolic model fit to human and animal experiments. ‘Kirklin – Clinical’ values are taken from Kirklin et al (2012) textbook, Figure 2-11 (‘X’ marks). ‘GARIX’ numbers are steady-state prediction of GARIX(7) model, i.e., without the age and weight terms and without temperature nonlinearity. ‘eGARIX’ columns show steady-state oxygen demand predictions from eGARIX(7) for different representative ages with weight for each age based on the prediction of a LOESS model regressing logarithm of weight on logarithm of age. Weights used are 3.0 kg (neonate), 4.8 kg (infant), 8.4 kg (toddler) and 26.5 kg (child).

| Temp | Kirklin | eGARIX |
| --- | --- | --- |

Table 1: Comparison of Kirklin and GARIX/eGARIX predictions for oxygen demand at different temperatures.
| (C) | Asymptotic | Clinical | neonate<br>(5-days) | infant<br>(3-month) | toddler<br>(1-year) | child<br>(8-years) |
| --- | --- | --- | --- | --- | --- | --- |
| 15 | 23 | NA | 6 | 9 | 11 | 10 |
| 20 | 35 | 25 | 11 | 18 | 20 | 18 |
| 25 | 55 | 45 | 21 | 33 | 38 | 34 |
| 30 | 86 | 65 | 37 | 59 | 69 | 61 |
| 37 | 161 | 120 | 46 | 73 | 85 | 76 |

### Oxygenation Gap Dynamics

Traditionally, it is assumed that the body extracts the oxygen it needs if supply is sufficient, allowing only under-oxygenation in static models. However, dynamically, over-oxygenation can occur in the short term. Figure 6 shows that immediately after a change in oxygen supply, the oxygenation gap (OG)—the percentage difference between VO_2_i and tVO_2_i—can be positive due to limited immediate adaptation in OER. Over time, OER adjusts, reducing the OG to zero in cases of surplus. Similarly, while a static view implies that under-oxygenation is only possible when OER exceeds a critical threshold (e.g., 0.60), our dynamic model predicts that, in the short-term, under-oxygenation can occur at much smaller OER levels. This is reflected in the near-symmetric shape of the OG line immediately after CI is changed (Figure 6B, light blue line).

**Figure 6.**
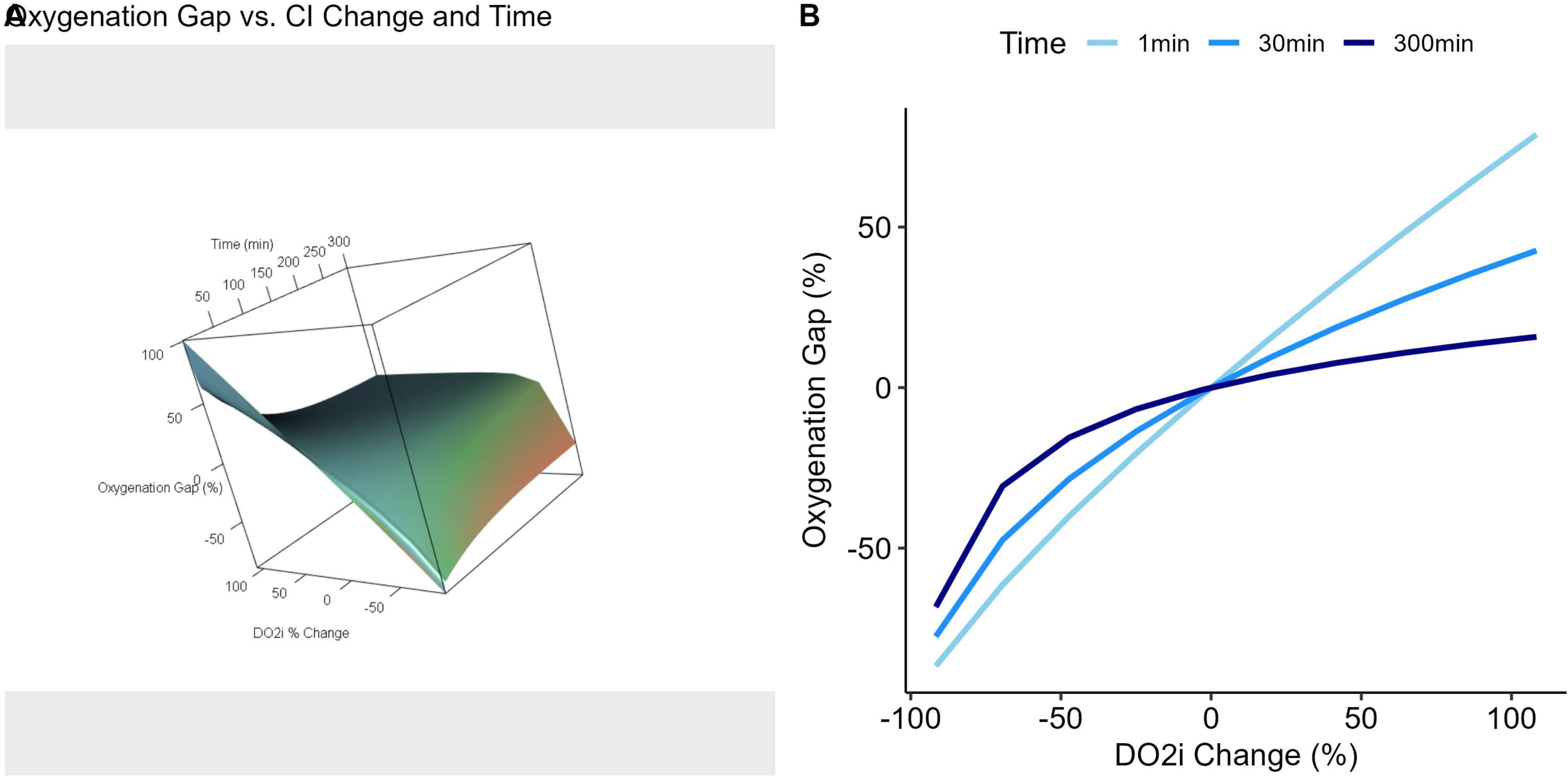
Short-Term vs. Long-Term Oxygenation Gap. Short-Term vs. Long-Term Oxygenation Gap (OG). Oxygenation gap (OG) as a function of % change in DO2i (via a step change in CI at t = 0), at different wait times, using deterministic simulation with a GARIX(7) model. State variables are log(CI) = 0.217, log(Hb) = 1.96, log(SaO2) = 4.6, temp = 20, logit(OER) = −1.42. Positive OGs indicate over-oxygenation and negative OGs reflect under-oxygenation. See text for exact definitions.

## Comment

### Oxygen Demand vs. Age and Weight

The eGARIX model reveals a nonlinear relationship between oxygen demand and age/weight, with oxygen demand per kilogram peaking around age 3 before declining. This pattern aligns closely with the Schofield BMR curve, providing a smooth and physiologically plausible model. These findings underscore the importance of individualizing oxygen delivery based on age and weight, as metabolic demands vary significantly during childhood growth and development.

We observed a progressively weaker OER response to haemoglobin changes in younger patients. In paediatric CPB, increasing haemoglobin often involves transfusion of stored red blood cells (RBCs), which are more commonly used due to circuit volume relative to patient size. Recent studies suggest that stored RBCs may unload oxygen more slowly, reducing O_2_ extraction [11]. Additionally, the oxygen dissociation curve may shift differently with stored RBCs due to storage-induced changes [12,13], affecting SaO_2_ dependence on PaO_2_.

### Dependence of Q10 on Temperature Regime

Allosteric enzymes, which regulate glycolysis, do not strictly follow van’t Hoff dependence on temperature and often exhibit sigmoid relationships [14]. Our eGARIX model shows that Q_10_ decreases significantly at higher temperatures, from 3.8 at 16°C to 1.2 at 37°C, suggesting a nonlinear dependence. This may be due to maintained enzymatic function and sufficient substrate availability under mild hypothermia, leading to decoupling between metabolic rate and temperature. As temperature decreases further, this independence diminishes, and higher Q_10_ values are observed. Further studies are needed to validate these findings and elucidate the underlying mechanisms.

### Limitations and Future Directions

Despite our model’s robustness, limitations exist. Our cohort includes only paediatric patients, potentially limiting generalizability to adults. Additionally, our model may not account for all patient-specific variables, such as genetics or comorbidities. Sparse low-temperature data limit conclusions about extreme hypothermia. Future studies should validate these findings in larger, diverse cohorts and explore temperature–oxygen demand mechanisms.

Combining neonates and teenagers, despite physiological differences, was necessary due to the low variable importance for the disequilibrium term group, which limited subgroup analyses. Building a single, global model maximized study power for steady-state analysis. More data are needed to differentiate age groups, particularly in cases like aortic arch repair where perfusion strategies differ.

## Conclusions

In conclusion, our extended model (eGARIX) offers novel insights into oxygen demand determinants in paediatric patients, emphasizing the nonlinear dependence on age, weight, and temperature. The observed relationships between OER and haemoglobin/SaO_2_ highlight the necessity for individualized oxygen delivery strategies, especially in neonates and infants. Our findings challenge assumptions like the fixed Q_10_ relationship, underscoring the value of flexible models in optimizing clinical decisions during CPB. Future research should validate these results in broader populations and refine oxygen demand management strategies, ultimately enhancing personalized care in critical settings.

## Data Availability

All data produced in the present study are available upon reasonable request to the authors.

## Declaration of Generative AI and AI-assisted technologies in the writing process

During the preparation of this work the author(s) used ChatGPT in order to edit and reorganise the manuscript. After using this tool/service, the author(s) reviewed and edited the content as needed and take(s) full responsibility for the content of the publication.

## References

1. Sharabiani, Mansour Taghavi Azar, et al. "A Dynamic Time-Series Model of Oxygen Consumption during Paediatric Cardiopulmonary Bypass." medRxiv (2024): 2024–03.

2. Kirklin, James K., and Eugene H. Blackstone. Kirklin/Barratt-Boyes Cardiac Surgery: Expert Consult-Online and Print (2-Volume Set). Elsevier Health Sciences, 2012.

3. Fox, Lawrence S., et al. "Relationship of whole body oxygen consumption to perfusion flow rate during hypothermic cardiopulmonary bypass." The Journal of Thoracic and Cardiovascular Surgery 83.2 (1982): 239–248.

4. Cheng, Huan-Chen, et al. "A study of oxygen consumption during extracorporeal circulation." ASAIO Journal 5.1 (1959): 273–278.

5. Paneth, Matthias, et al. "Physiologic studies upon prolonged cardiopulmonary by-pass with the pump-oxygenator with particular reference to (1) acid-base balance,(2) siphon caval drainage." Journal of Thoracic Surgery 34.5 (1957): 570–579.

6. Starr, Albert. "Oxygen consumption during cardiopulmonary bypass." The Journal of Thoracic and Cardiovascular Surgery 38.1 (1959): 46–56.

7. Rogers, Libby, et al. "Improving risk adjustment for mortality after pediatric cardiac surgery: the UK PRAiS2 model." The Annals of thoracic surgery 104.1 (2017): 211–219.

8. Schofield, William N. "Predicting basal metabolic rate, new standards and review of previous work." Human nutrition. Clinical nutrition 39 (1985): 5–41.

9. Khwaja A. KDIGO clinical practice guidelines for acute kidney injury. Nephron Clin Pract 2012;120:c179–184. doi: 10.1159/000339789

10. Naik SK, Elliott MJ. Ultrafiltration and paediatric cardiopulmonary bypass. Perfusion 1993;8:101–112. doi: 10.1177/026765919300800114

11. Dumbill R, Rabcuka J, Fallon J, et al. Impaired O2 unloading from stored blood results in diffusion-limited O2 release at tissues: evidence from human kidneys. Blood 2024;143:721–733. doi: 10.1182/blood.2023022385

12. Bennett-Guerrero E, Veldman TH, Doctor A, et al. Evolution of adverse changes in stored RBCs. Proc Natl Acad Sci U S A 2007;104:17063–17068. doi: 10.1073/pnas.0708160104

13. Yoshida T, Prudent M, D’Alessandro A. Red blood cell storage lesion: causes and potential clinical consequences. Blood Transfus 2019;17:27–52. doi: 10.2450/2019.0217-18

14. Lenzen S. A fresh view of glycolysis and glucokinase regulation: history and current status. J Biol Chem 2014;289:12189–12194. doi: 10.1074/jbc.R114.557314

15. Hall, John E. Guyton and Hall Textbook of Medical Physiology, Jordanian Edition E-Book. Elsevier Health Sciences, 2016.

