## Supplementary Material for "Mathematical Modelling of Oxygenation Dynamics using High-Resolution Perfusion Data – Part 2: Physiological Insights"

**Brief Title**: Paediatric CPB Oxygenation Dynamics

**Authors**: Mansour T. A. Sharabiani, PhD[a†], Alireza S. Mahani, PhD[b†], Richard W. Issitt, DClinP[c,d,e†], Yadav Srinivasan, MD[f], Alex Bottle, PhD[a], Serban Stoica, FRCS MD[g].

**Affiliations**:

a. School of Public Health, Imperial College, London, United Kingdom

b. Statman Solution Ltd., London, United Kingdom

c. Centre for Heart Failure, Transplantation and Extracorporeal Support, UCL Institute for Cardiovascular Science, UCL, London, United Kingdom

d. Perfusion Department, Great Ormond Street Hospital for Children, London, United Kingdom

e. Data Research, Innovation and Virtual Environments Unit, Great Ormond Street Hospital for Children, London, United Kingdom

f. Cardiac Surgery Department, Great Ormond Street Hospital for Children, London, United Kingdom

g. Cardiac Surgery Department, Bristol Royal Children's Hospital, Bristol, United Kingdom

† These authors contributed equally to this work.

The authors have no conflicts of interest to disclose.

**Corresponding Author**:

Dr Mansour T A Sharabiani

School of Public Health, Imperial College London

London, United Kingdom, W12 0BZ

**Corresponding Co-Author**:

Dr Richard Issitt

Centre for Heart Failure, Transplantation and Extracorporeal Support, Research Department of Children's Cardiovascular Disease, UCL Institute of Cardiovascular Science, London, United Kingdom.

### Supplementary Materials

#### A: Nonstandard Abbreviations and Acronyms

- AKI: acute kidney injury
- ATG: autoregressive term group
- BMR: Basal Metabolic Rate
- BSA: body surface area ($m^2$)
- CI: cardiac index (blood flow indexed to BSA; L/min/$m^2$)
- CPB: cardiopulmonary bypass
- CV: cross-validation
- DHCA: deep hypothermic circulatory arrest
- $DO_2$: oxygen delivery (mL/min)
- $DO_2i$: oxygen delivery, indexed to BSA (mL/min/$m^2$)

- EHR: electronic health record
- ETG: equilibrium term group
- GARIX: A global autoregressive integrated (time-series) model with exogenous variables, plus a disequilibrium term group
- eGARIX: Extended GARIX, i.e., GARIX with additional terms in the ETG for patient age, weight and the interaction of age and weight, and replacing the linear term for temperature with a second-order spline.
- GDP: goal-directed perfusion
- Hb: haemoglobin content of blood (g/dL)
- i.i.d: independent and identically distributed
- LOESS: Locally estimated scatterplot smoothing
- ML: machine learning
- NICOR: national institute for cardiovascular outcomes research
- OER: oxygen extraction ratio
- OG: oxygenation gap
- OOS: out-of-sample
- $PaO_2$: partial pressure of oxygen (kPa)
- PRAiS2: partial risk adjustment in surgery model
- PVI: permutation-based variable importance
- Q10: multiplicative increase in the resting metabolic rate for every 10℃ increase in body temperature
- RBCs: red blood cells
- RMR: resting metabolic rate
- $SaO_2$: oxygen saturation of arterial blood (%)
- $SvO_2$: oxygen saturation of mixed venous blood (%)
- TG: term group

- $tVO_2i$: target (indexed) oxygen consumption (indexed oxygen demand; mL/min/$m^2$)
- $tDO_2i$: threshold (indexed) oxygen delivery (mL/min/$m^2$)
- VI: variable importance
- $VO_2$: oxygen consumption (mL/min)
- $VO_2i$: oxygen consumption, indexed to BSA (mL/min/$m^2$)
- XTG: Exogenous Term Group

### B: Methods

#### Data Collection

Routine clinical information was extracted from the institution's Electronic Health Record (EHR) system using a custom structured query language script. These data included demographic information, laboratory results, intraoperative, CPB, and medication administration data, and intensive care requirements. Comorbidities (including risk scores), and outcomes were defined using Association for European Paediatric and Congenital Cardiology coding as part of the institution's routine NICOR data submissions. Intraoperative data from the heart-lung and anaesthetic machines were captured every minute throughout surgery. Preoperative risk estimation was based on cardiac and non-cardiac preoperative risk factors using the Partial Risk Adjustment in Surgery (PRAiS2) model [7]. Baseline serum creatinine levels were taken within the 24 hours preceding surgery. Intraoperative blood biochemistry (including haemoglobin, arterial oxygen partial pressure and saturation, and mixed-venous oxygen saturation) was measured continuously using an optical fluorescence and reflectance system (CDI550, Terumo, Leuven, Belgium), calibrated every 20 minutes using a cassette-based blood gas analyser (ABL90 Flex Plus, Radiometer, Copenhagen, Denmark). HLM variables were automatically recorded in one-minute intervals via a bidirectional connection into the Institutional EHR system (Axon, Philips Capsule, Paris, France). Postoperative AKI was calculated according to the Kidney Disease Improving Global Outcomes (KDIGO) criteria based on the patients' serum creatinine change and urine levels within the first 48 hours following surgery [8]. Patients were assigned the highest AKI grouping on the basis of these two criteria. The final KDIGO score for each patient is an integer between 0 (no AKI) and 3 (severe AKI).

#### Anaesthesia

Anaesthesia was induced by inhalation of sevoflurane in oxygen and, after induction, fentanyl 5 mg/kg and pancuronium100 mg/kg were given; maintenance was with isoflurane 1.0% in oxygen and air. After tracheal intubation, arterial and central venous lines were inserted. Further incremental doses of fentanyl, up to 25 mg/kg, were given during the procedure.

#### CPB

The CPB circuit consisted of FX oxygenators with integrated arterial line filter and hard-shell venous reservoir (Terumo). The Stöcket S5 (Stöcket, LivaNova, Munich, Germany) heart-lung machine with pole-mounted roller pumps was used with a 3T heater-cooler (Stöcket). The total base prime volume was 330 - 1000 mL depending on circuit size. When the predicted initial CPB haematocrit was <27% the prime consisted of packed red blood cells of the patient's blood group (120-200mL), human albumin solution (50-150mL), heparin 1000 U/mL, 1.5 mL (Heparin Sodium; Wockhart, Wrexham, UK). Blood primed circuits were then washed with 1000 mL of balanced crystalloid solution (Plasmalyte 7.4; Baxter, Thetford, UK) by performing prebypass ultrafiltration, carried out as previously described, to ensure an initial on-CPB haematocrit of 30% [9]. Biochemical compatibility was then attained using sodium bicarbonate as a buffering agent. Patients requiring a clear prime received crystalloid and human albumin solution, with 10 mL sodium bicarbonate and 2 to 5 mL heparin. Myocardial protection was achieved with 30 mL/kg of cold blood cardioplegia (4:1 blood: cardioplegia to a final potassium concentration of 20 mmol/L) of St Thomas's Solution (IVEX Pharmaceuticals, Larne, Northern Ireland). During the rewarming phase of CPB, a gradient no greater than 10°C between the patient's oesophageal temperature probe and the heater-cooler (maximum arterial blood temperature 37.5°C) was maintained until a maximum oesophageal temperature of 36°C was achieved.

### C: OER Dynamics by Age Group – Remaining Variables

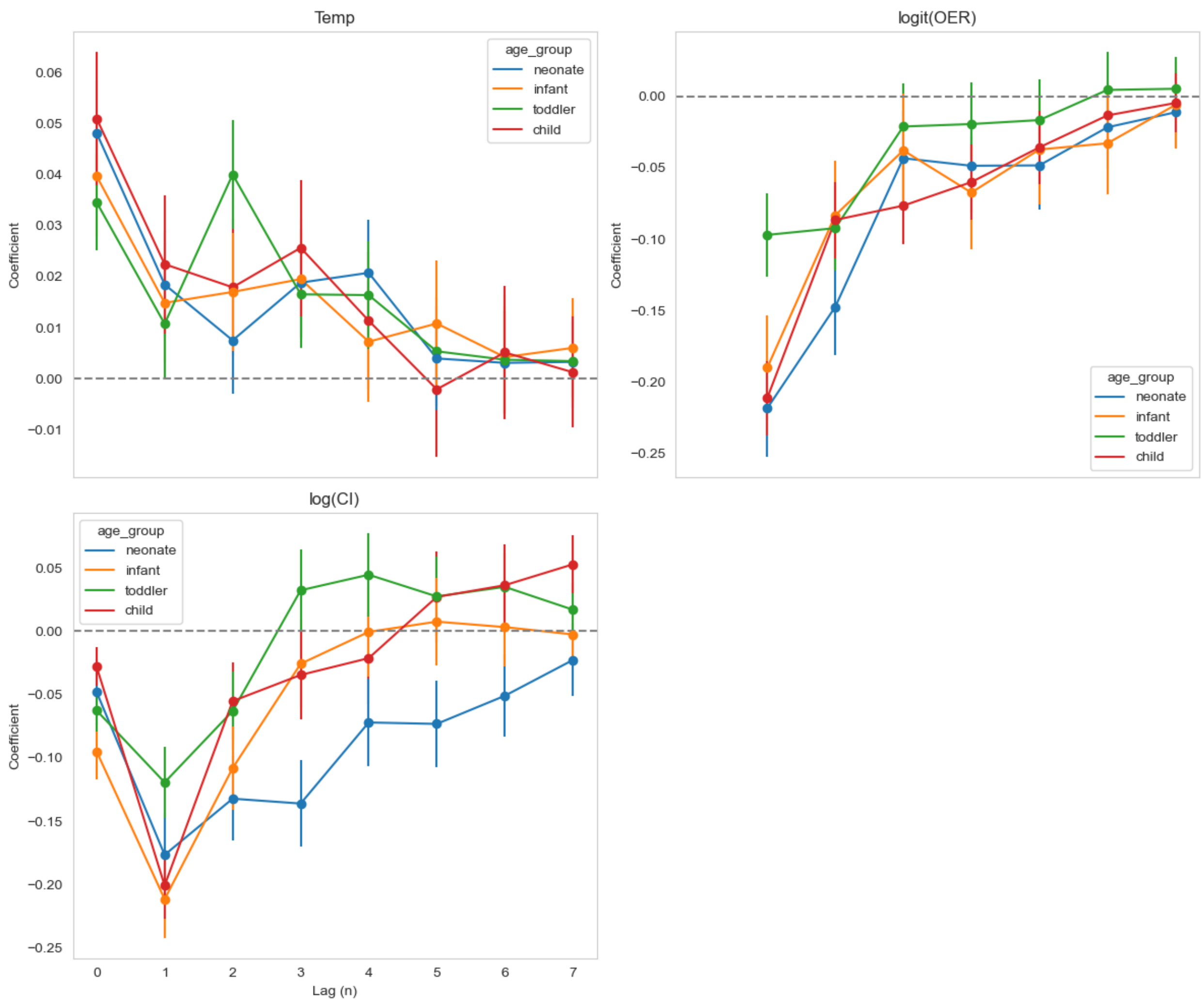


Figure C.1: Coefficients of change terms – as a function of lag $(n)$ – in eGARIX for $T$, $logit(OER)$ and $\log CI$, stratified by age group.

### D: Fox82 Human Experiments and the Kirklin Equation

In this section, we briefly describe the experimental setup of Fox82 as well their synthesis of the human experiments they conducted along with previous experiments on animals. For details, see Fox et al (1982).

#### Fox82 Experiments

A total of 17 adults (15 males, 2 females) were included whose age was 57+/-10 years (mean +/- std). Mean body temperature during recordings was 21.3C. Initial $CI$ was 2.2 L/min/m2. Mean venous $Hb$ was 7.1. $SaO_2$ was not given, but we assumed a value of 99%, consistent with their assertion that 100% oxygen (no CO2) was used for ventilator oxygenation. After an initial stabilisation period, a randomly-shuffled sequence of five nominal $CI$'s (0.25, 0.5, 1.0, 1.5 and 2.0 L/min/m2) were used with each patient, holding each level for 10 minutes. After device recalibration, actual flows were estimated to be 0.25 +/- 0.084, 0.54 +/- 0.101, 1.02 +/- 0.107, 1.56 +/- 0.129 and 2.08 +/- 0.180. We used these mean and std values to draw $CI$'s for our simulations. Due to surgical constraints, the average number of 10-min episodes used with each patient was 4 (rather than 5).

#### Kirklin Equation

Kirklin et al (2012) combine the hyperbolic expression for the dependence of oxygen consumption on cardiac flow with the Van 't Hoff expression for the dependence of oxygen demand on temperature to arrive at the following equation for oxygen consumption ($VO_2i$) as a function of cardiac index ($CI$) and temperature ($Temp$):

$$\frac{1}{VO_2i} = 0.168 \times 10^{-0.0387\,Temp} + \frac{0.0378}{CI} \times 10^{-0.0253\,Temp} \qquad \text{(D.1)}$$

The numbers are based on fitting the equation to the human experiments of Fox82 done at 20C with the animal experiments of [4-6] done at 37C. Equation D.1 implies a $Q_{10}$ for oxygen demand (asymptotic oxygen consumption at infinite $CI$) of 2.4.

Evaluating the above equation at $Temp = 20$ produces the following hyperbolic equation, which is what Fox82 reported in their paper:

$$VO_2 i \; = 35 \; \times CI \; / \; (0.42 \; + \; CI) \qquad \text{(D.2)}$$

### GARIX vs. Fox82 – Hyperbolic Parameters

As shown in the table below, the estimated parameters of the hyperbolic fit by Fox82 fall within the confidence intervals from our simulation of Fox82 experiments using GARIX. Same applies to the R-squared of the fitted hyperbolic models.

| | **GARIX(7)** | | **Fox82** |
|---|---|---|---|
| **Parameter** | **estimate** | **95% CI** | **estimate** |
| **R-squared** | 72.1% | 48.4% - 92.9% | 65% |
| **a** | 30.8 | 21.3 – 46.6 | 35 |
| **b** | 0.84 | 0.39 - 1.54 | 0.42 |

Table D.1: Comparison of R-squared and fit parameters from GARIX(7) simulations mimicking experiments of Fox82 [2]. See Supplementary Material D for model specification in Fox82 and their definitions of ‘a’ and ‘b’. Confidence intervals are based on 100 runs.
